# Hot Spring Residency and Disease Association: a Crossover Gene-Environment Interaction (GxE) Study in Taiwan

**DOI:** 10.1101/2024.07.29.24311167

**Authors:** Hsin-Yu Wu, Kao-Jung Chang, Wei Chiu, Ching-Yun Wang, Yu-Tien Hsu, Yuan-Chih Wen, Pin-Hsuan Chiang, Yu-Hsiang Chen, He-Jhen Dai, Chia-Hsin Lu, Yi-Cheng Chen, Han-Ying Tsai, Yu-Chun Chen, Chih-Hung Hsu, Ai-Ru Hsieh, Shih-Hwa Chiou, Yi-Ping Yang, Chih-Chien Hsu

**Affiliations:** Department of Medical Research, Taipei Veterans General Hospital, No.201, Sec. 2, Shipai Rd., Beitou District, Taipei 11217, Taiwan; School of Medicine, National Yang Ming Chiao Tung University, No. 155, Sec. 2, Linong St. Beitou Dist., Taipei 112304, Taiwan; Institute of Clinical Medicine, National Yang Ming Chiao Tung University, No. 155, Sec. 2, Linong St. Beitou Dist., Taipei 112304, Taiwan; Department of Ophthalmology, Taipei Veterans General Hospital, No.201, Sec. 2, Shipai Rd., Beitou District, Taipei 11217, Taiwan; Department of Medical Education, Taipei Veterans General Hospital, No.201, Sec. 2, Shipai Rd., Beitou District, Taipei 11217, Taiwan; Department of Medical Education, Taichung Veterans General Hospital, No. 1650, Taiwan Boulevard Sect. 4, Taichung 407219, Taiwan; Department of Social & Behavioral Sciences, Harvard T.H. Chan School of Public Health, 677 Huntington Ave, Boston, MA 02115, USA; Department of Occupational Medicine and Clinical Toxicology, Taipei Veterans General Hospital, No.201, Sec. 2, Shipai Rd., Beitou District, Taipei 11217, Taiwan; Institute of Environmental and Occupational Health Sciences, National Yang Ming Chiao Tung University, No. 155, Sec. 2, Linong St. Beitou Dist., Taipei 112304, Taiwan; Big Data Center, Taipei Veterans General Hospital, No.201, Sec. 2, Shipai Rd., Beitou District, Taipei 11217, Taiwan; Department of Statistics, Tamkang University, No.151, Yingzhuan Rd., Tamsui Dist., New Taipei 251301, Taiwan; Institute of Hospital and Health Care Administration, National Yang Ming Chiao Tung University, No. 155, Sec. 2, Linong St. Beitou Dist., Taipei 112304, Taiwan; Department of Family Medicine, Taipei Veterans General Hospital, No.201, Sec. 2, Shipai Rd., Beitou District, Taipei 11217, Taiwan; Women’s Hospital, The Fourth Affiliated Hospital, and Department of Environmental Medicine, Zhejiang University School of Medicine, No. 866, Yuhangtang Rd, Hangzhou 310058, China; Zhejiang University School of Medicine, Hangzhou 310006 Institute of Genetics, International School of Medicine, Zhejiang University, No. 866, Yuhangtang Rd, Hangzhou 310058, China; Institute of Pharmacology, National Yang Ming Chiao Tung University, No. 155, Sec. 2, Linong St. Beitou Dist., Taipei 112304, Taiwan; Institute of Food Safety and Health Risk Assessment, School of Pharmaceutical Sciences, National Yang Ming Chiao Tung University, No. 155, Sec. 2, Linong St. Beitou Dist., Taipei 112304, Taiwan

**Keywords:** Gene-environment association, Hot spring, Dry eye disease, Valvular heart disease, Genome-wide association study, Taiwan biobank

## Abstract

**Background:** The advent of genetic biobanking has powered gene-environment interaction (GxE) studies in various disease contexts. Therefore, we aimed to discover novel GxE effects that address hot spring residency as a risk to inconspicuous disease association.

**Methods:** A complete genetic and demographic registry comprising 129,451 individuals was obtained from Taiwan Biobank (TWB). Geographical disease prevalence was analyzed to identify putative disease association with hot-spring residency, multivariable regression and logistic regression were rechecked to exclude socioeconomic confounders in geographical-disease association. Genome-wide association study (GWAS), gene ontology (GO), and protein-protein interaction (PPI) analysis identified predisposing genetic factors among hotspring-associated diseases. Lastly, a polygenic risk score (PRS) model was formulated to stratify environmental susceptibility in accord to their genetic predisposition.

**Results:** After socioeconomic covariate adjustment, prevalence of dry eye disease (DED) and valvular heart disease (VHD) was significantly associated with hot spring distribution. Through single nucleotide polymorphisms (SNPs) discovery and subsequent PPI pathway aggregation, CDKL2 and BMPR2 kinase pathways were significantly enriched in hot-spring specific DED and VHD functional SNPs. Notably, PRS predicted disease well in hot spring regions (PRS_DED_: AUC=0.9168; PRS_VHD_ AUC=0.8163). Hot spring and discovered SNPs contributed to crossover GxE effect on both DED (relative risk (RR)_G+E-_=0.99; RR_G+E+_=0.35; RR_G+E+_=2.04) and VHD (RR_G+E-_=0.99; RR_G+E+_=0.49; RR_G+E+_=2.01).

**Conclusion:** We identified hot-spring exposure as a modifiable risk in the PRS predicted GxE context of DED and VHD.

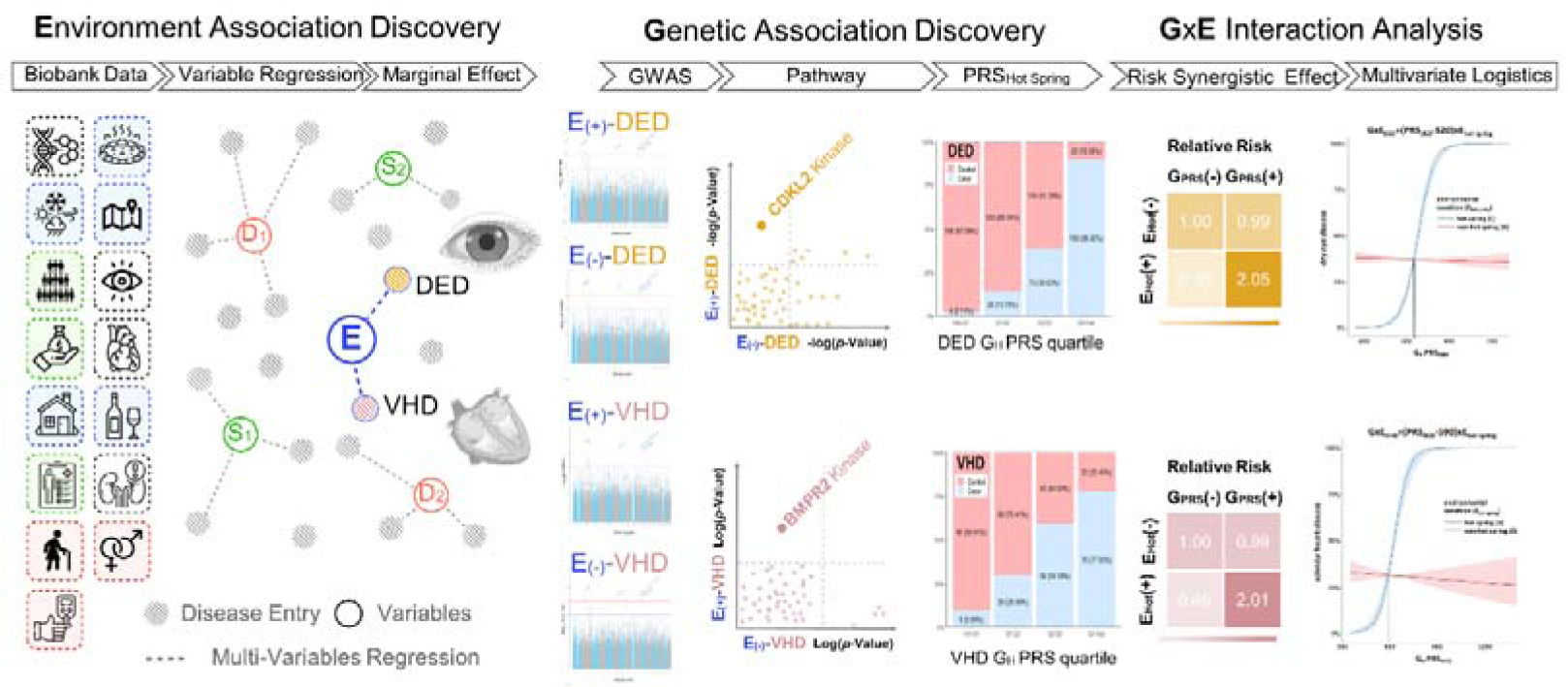

## 1. Introduction

Gene-environment interaction (GxE) embodies a pathogenic context in which the impact of environmental exposures modifies our susceptibility to genetic mutations (Hunter 2005). Although the GxE risk addition effects are reported to be potent and widely existed, GxE effects are often hard to discover and therefore overlooked from the scope of general pathophysiology. To this end, the advent of big data and the genetic biobanking era has enabled diverse GxE studies in the development of complex disease (McAllister et al. 2017). While environmental stimuli such as ultraviolet radiation, air pollutants (such as PM2.5, CO, and sulfates), and environmental toxins have been extensively studied in the context of GxE mechanisms (Virolainen et al. 2022), scant attention has been paid to the environmental effect of hot spring exposure—which allegedly can bring forth health benefits, hence we set goal to understand the hot-spring related GxE interactions.

Throughout ancient cultures ranging across the empire of Rome, China, Japan, and the islanders of Austronesian, hot springs bathing was deemed to improve health, gain cosmetic benefit, and expand life. Recent scientific studies have gathered complementary evidence indicating that hot spring bathing could prevent diseases, alleviate musculoskeletal discomfort, modulate nerve and endocrine systems, and promote immunity against chronic diseases (such as whip syndrome, rheumatoid arthritis (RA), cancer, hemorrhoids, and skin diseases) (Bidonde et al. 2014; Verhagen et al. 2015). Moreover, Onsen therapy was certified by the Japanese Society for Complementary and Alternative Medicine (JCAM) (Serbulea and Payyappallimana 2012), and was subsidized by health insurance in European countries (such as France, Germany, and Italy).

Hot spring is a geothermal heated water body that exerts indirect health influences through remodeling the local atmosphere and local hydrology. Bioactive gaseous substances such as H_2_S generally range between 1.0 and 3.0 μg/m^3^ in urban areas, whereas the H_2_S level spiked as high as 1500 μg/m^3^ in the residential vicinity of geothermal (Gorini et al. 2020). The exact effect of these bioactive geothermal-released substances was not fully understood, but independent pilot studies had reported positive association between H_2_S exposure and hospitalization events related to circulatory systems (Standardized Hospitalization Ratios (SHR)=1.04; 95% CI=1.01–1.07) (Nuvolone et al. 2019). Consistently, another geothermal epidemiology study in Italy also showed that the increased H_2_S exposure in men could result in higher cardiovascular mortality rates (odds ratio (OR)=1.22, 90% CI=1.03-1.44) (Nuvolone et al. 2020). Furthermore, a cohort study in Rotorua (New Zealand) showed that dwelling in geothermal emission areas was associated with increased malignancy rate and a 1.26 (95% CI=1.14–1.38) standardized incidence ratio (SIR) for cataract.(Bates et al. 1998)

While hot spring bathing was reported to bring folk-healing benefits to musculoskeletal diseases, hot spring residency was also linked to cardiovascular threats, therefore the health influence of hot spring exposure remains controversial to date.

In this study, we set goal to investigate the crossover effect between hot spring residency and genetic predisposition. To be specific, we performed a three-component study that included environmental association discovery, variant-disease association discovery, and GxE interaction analysis to delineate the environmental effects of hot spring exposure on orthopedic, ophthalmic, and cardiopulmonary diseases.

## 2. Materials and Methods

### 2.1. Study Design

This is a three-component study that includes environmental association discovery (demographic analysis and logistic regression model), variant-disease association discovery (exposure stratified GWAS, protein-protein interaction network analysis and polygenic risk score), and the GxE interaction analysis (crossover GxE effects and multivariate accumulative analysis).

### 2.2. Study Population

Nation-wide TWB1 and TWB2 (https://www.twbiobank.org.tw/) participant registry was obtained. The 129451 individuals were enrolled from the general population by TWB1 and TWB2 between 2016 to 2023. At the time of entering the cohort, all participants were cancer-free and were obligated to complete a structured questionnaire that documented: sociodemographic characteristics, lifestyle, dietary habits, environmental exposures, and major medical histories (Fan et al. 2008). The disease case labels in TWB were self-reported, and control cases were designated by disease-free individuals that were above 60 years old.

### 2.3. Outcomes of Interest

The primary outcomes of interest of this study was set on hot-spring related orthopedic diseases (osteoporosis, arthritis, gout), ophthalmic diseases (cataracts, glaucoma, dry eye disease; DED, retinal detachment; RD, floaters, blindness, color vision deficiency; CVD and age-related macular degeneration; AMD), cardiopulmonary diseases (asthma, valvular heart disease; VHD, coronary artery disease; CAD, arrhythmia, cardiomyopathy, congenital heart disease, hyperlipidemia, hypertension, stroke, diabetes).

### 2.4. Exposure of Interest

The exposure of interest in this study includes hot spring area residency and polygenetic underlying with adjustment of socioeconomic confounder.

For which we divided hot-spring and non-hot-spring residential areas by the proclaiment of the National water resource department. The hot spring definitions were: water body heated above 30 celsius with total dissolved solid >500 mg/L and meeting one additional minor criterions in category (I-a) HCO ^-^ >250 mg/L (I-b) SO ^2-^ >250 mg/L (I-c) Cl^-^ and other halides > 250 mg/L, or (II-a) free CO_2_ >250 mg/L (II-b) total sulfide >0.1 mg/L (II-c) total Fe^2+^ and total Fe^3+^ >10 mg/L (II-d) Radium >10^-8^ curie/L. The polygenetic underlying was primarily assessed through variant-disease association (VDA), which identified disease-associated SNPs through GWAS. Secondly, independent polygenic risk score (PRS) models were established based upon the GWAS summarized SNP lists, whereon each PRS predicts the corresponding disease risk for hot spring residents and non-hot spring residents. The functional PRS SNP genes were further analyzed with gene ontology (GO), co-expressed signaling pathways, and protein-protein interaction (PPI) network analysis.

### 2.5. Adjusted Covariates for Hot spring Exposure and Disease Outcome

We first performed descriptive analysis. Continuous data are presented as means with standard deviation, and categorical data are presented as proportions. Our study used the Student’s t-tests (normally distributed data) or Wilcoxon tests (non-normally distributed data) for continuous variables and chi-squared tests or fisher exact test for categorical variables between two groups (hot spring residents vs controls).

To find hot spring exposure related diseases, odds ratio (OR) and logistic regression were conducted to detect disease association with hot spring residency, a subsequent multivariate analysis was performed to solidify disease association under the consideration of covariate adjustments. Since hot springs are mainly distributed in townships with deviated sex ratios, imbalanced age composition, sparse medical institutes, and less income, these socioeconomic and demographic covariates may confound with the relationship between hot spring exposure (E) and the disease outcome (D) (Supplementary Table 1 and Figure 1).

**Figure 1:**
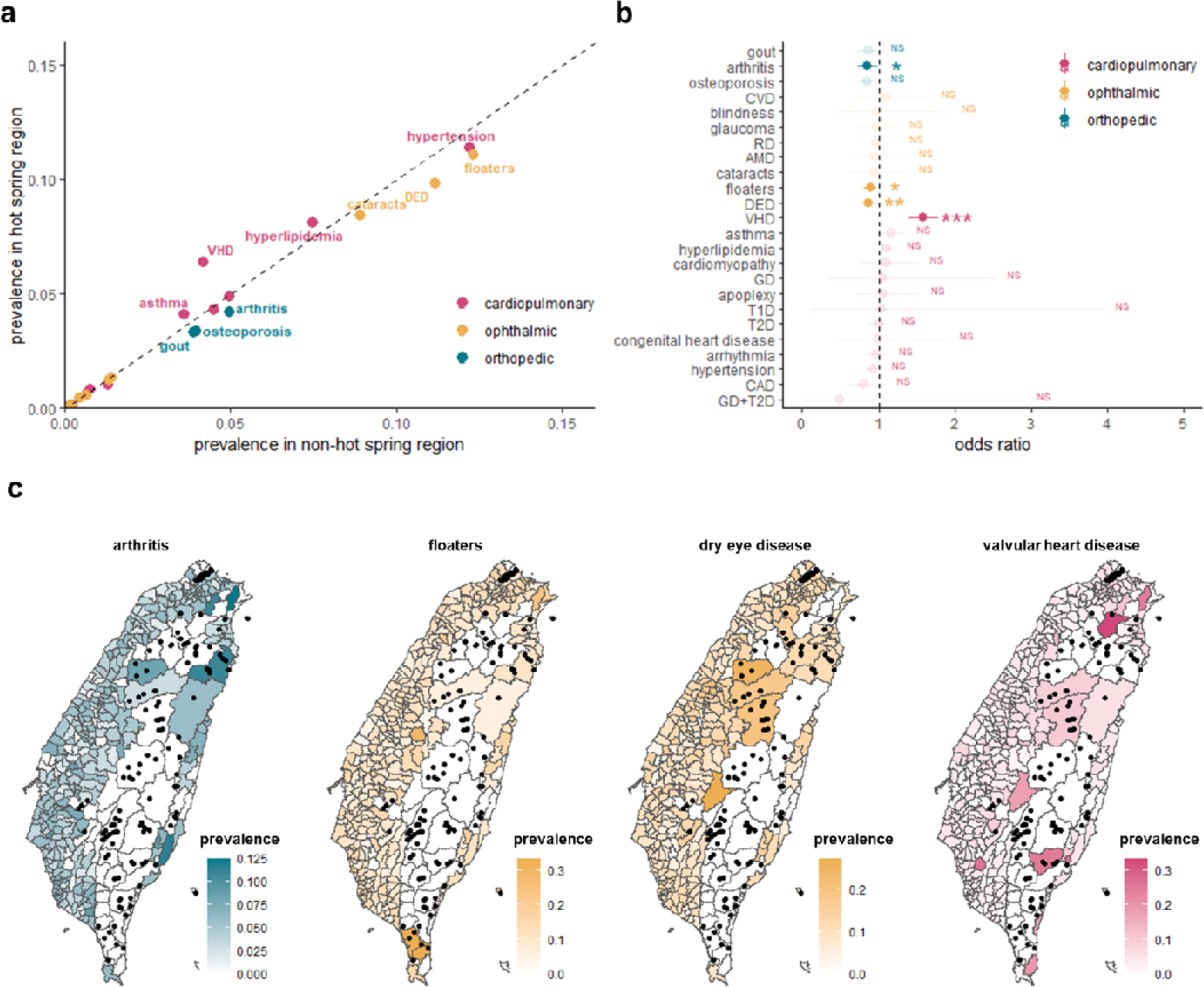
Demographic Analysis Finding Environmental (E) Effect. (a) Comparing the disease prevalence between hot spring region and non-hot spring region. The dashed line indicated the same disease prevalence in both regions. (b) Odds ratio (OR) indicating associations between hot spring geology and disease prevalence with 95% confidence interval (CI). The dashed line indicated OR=1.0 (c) Prevalence distribution of hot spring-related disease. The dots indicated the location of the hot spring outcrop. DED, dry eye disease; RD, retinal detachment; CVD, color vision deficiency; AMD, age-related macular degeneration; VHD, valvular heart disease; CAD, coronary artery disease; GD, gestational diabetes (GD); T1D, type 1 diabetes; and T2D, type 2 diabetes.

To specify the data source of the adjusted covariates: the demographic covariates, including age, sex ratio and total resident counts, were recorded from the Directorate-General of Budget, Accounting and Statistics. Total comprehensive income was collected from the released tolls of the Ministry of Finance (https://www.mof.gov.tw/). Accessible medical resources were estimated by health insurance index (sum of health insurance benefits spent among all the hospitals in the township) and medical visit index (sum of annual medical visits among all the hospitals in the township) from the auditory releases of the Ministry of Health and Welfare (https://data.gov.tw/dataset/39280). Continuous variables that did not follow a normal distribution were provided with the mean±standard deviation (SD) and quartile-based (min, Q1, Q2, Q3, and max) categorical conversion. The first category was pinned as reference, then the effect of covariates was evaluated by univariable and multivariable logistic regression models.

Univariate logistic regression analyses were used to show the significant and independent role of the different variables in determining outcome (including arthritis, floaters, DED, and VHD) effects. Multivariate logistic regression analyses were used to control for confounding variables. All variables were then used for a multivariate logistic regression model with stepwise selection for constructing prediction models. Since in logistic regressions, estimated coefficients cannot be interpreted as a measure of the contribution of the effect, we have also calculated marginal effects. The marginal effect reflects the association between a small change in a variable and the alteration in probabilities across each outcome. These marginal effects were computed while keeping all other variables constant at the mean of the entire sample. Subsequently, multivariable logistic regression analyses were employed to investigate potential gene-environment interactions, utilizing the most suitable genetic model available. The predictive performance of the constructed predictive models was evaluated using sensitivity, specificity, and areas under the receiver operating characteristic (ROC) curves, the AUC value. All statistical analyses were performed using R version 4.1.1. The tests were 2 tailed, and *p*-value <0.05 was taken as statistically significant.

### 2.6. Variant-disease Association Discovery

All SNPs discoveries were performed on PLINK (version 1.9 downloaded from http://zzz.bwh.harvard.edu/plink/). SNPs quality control was conducted with the following criteria: (i) SNPs missing in more than 2% of participants (ii) individuals missing more than 5% SNP data (iii) SNPs with minor allele frequency (MAF) under 0.05 (iv) SNPs deviating from Hardy–Weinberg equilibrium (*p*-value <0.05) (v) individuals with identity by descent (IBD) over 0.125.

GWAS identified disease-specific SNPs for populations living in townships with and without hot springs. SNPs were further screened by pairwise squared correlation (r^2^) less than 0.01 that linkage disequilibrium (LD) was estimated with a window size of 4000 bp. Functional SNP genes were labeled as G_A_ for significant SNPs from all populations, G_N_ for significant SNPs from non-hot spring populations; G_H_ for significant SNPs from hot spring populations, and G_h_ indicated hot spring-specific functional SNP genes G_H_ that did not overlap in G_A_ and G_N_. Furthermore, based upon the GWAS summarized SNP lists, each PRS predicts the corresponding disease risk for hot spring residents and non-hot spring residents.

The functional PRS SNP genes were further analyzed on Enrichr (Chen et al. 2013; Kuleshov et al. 2016; Xie et al. 2021) to attribute gene ontology (GO) and the ARCHS4 Kinases Coexp library was applied to identify co-expressed signaling pathways. protein-protein interaction (PPI) network analysis of G_h_ was further evaluated in the Search Tool for Retrieval of Interacting Genes/Proteins (STRING version 11.5 download from http://string-db.org/) (Szklarczyk et al. 2018) with a confidence level of 0.400 (medium) based on the find of: experiments, databases, co-expression, neighborhood, gene function, and co-occurrence. Then, the activated pathways in the selected PPI network were further calculated with the Reactome 2022 database.

### 2.7. GxE Interaction Analysis

Crossover GxE effects between environmental conditions and individual genetic variants were assessed by relative risk (RR) with hot spring and PRS score as binary variables: living in township with/without hot spring and high/low PRS score.

Visualized by nomogram, multivariate accumulative analysis based on logistic regression model estimated predictability of confounders (sex) and GxE effect. Decision curve analysis (DCA) was performed to evaluate a clinical “net benefit” for nomogram prediction models in comparison to default strategies of treating all or no patients. Net benefit is calculated across a range of threshold probabilities, defined as the minimum disease risk at which further intervention would be warranted.

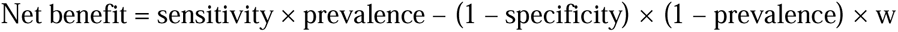

where w is the odds at the threshold probability.(Vickers et al. 2019)

## 3. Results

### 3.1. Finding Hot Spring Associated Disease

This study included 129,451 participants with 4517 individuals dwelled in the townships with hot spring outcrops; their demographic characteristics were detailed as in Table 1.

**Table 1:** Demographic Information of the Participants.

|  | **Hot spring residents**<br>**(n=4517)** | **Control**<br>**(n=124934)** | **Number (%)** | ***p*-value** |
| --- | --- | --- | --- | --- |
| **Age (years)** | 48.56±10.94 | 49.76±10.96 | 129451 (100.0) | <0.001 |
| **Body mass index(kg/m<sup>2</sup>)** | 24.03±3.40 | 24.00±3.40 | 126783 (97.9) | 0.682 |
| **Water daily intake (ml)** | 1286.80±633.15 | 1255.12±685.91 | 21735 (16.8) | 0.07 |
| **Sex** |  |  |  |  |
| Male (%) | 1658 (36.7) | 45066 (36.1) | 129451 (100.0) | 0.392 |
| Female (%) | 2859 (63.3) | 79868 (63.9) |  |  |
| **Alcoholism** |  |  |  |  |
| No or seldom (%) | 4122 (91.3) | 114134 (91.4) | 129338 (99.9) | 0.578 |
| Abstain (%) | 111 (2.5) | 3268 (2.6) |  |  |
| Yes (%) | 282 (6.2) | 7421 (5.9) |  |  |
| **Smoking experience** |  |  |  |  |
| Yes (%) | 3213 (71.2) | 90795 (72.7) | 129404 (100.0) | 0.025 |
| No (%) | 1301 (28.8) | 34095 (27.3) |  |  |
| **Water origin** |  |  |  |  |
| Tap water (%) | 77 (1.7) | 2716 (2.2) | 21958 (17.0) | 0.018 |
| Shallow well water (%) | 0 (0.0) | 13 (0.0) |  |  |
| Deep well water (%) | 9 (0.2) | 126 (0.1) |  |  |
| Mineral water (%) | 35 (0.8) | 1125 (0.9) |  |  |
| Filtered water (%) | 390 (8.6) | 15934 (12.8) |  |  |
| Other (%) | 49 (1.1) | 1484 (1.2) |  |  |

Among the included disease spectrum, four diseases (arthritis, floaters, dry eye disease and valvular heart disease) showed higher prevalence in hot-spring areas. (Figure 1a and 1b). However, the coefficient of determination of these diseases were not significantly associated with hot spring distribution (Figure 1c; R^2^=4.40% in arthritis, 2.51% in floaters, 1.54% in DED, and 4.53% in VHD)

After univariate and multivariate logistic regression, DED (multivariable: OR=0.78, 95% CI=0.65-0.92; stepwise: OR=0.81, 95% CI=0.679-0.961) and VHD (multivariable: OR=1.56, 95% CI=1.20-2.03; stepwise: OR=1.52, 95% CI=1.20-1.91) stood out as the two remaining hot spring-associated diseases (*p*-value<0.05); whereas the hot spring association with arthritis (*p*-value=0.0589 for multivariable; *p*-value=0.1496 for stepwise) and floaters (*p*-value=0.6807 for multivariable; *p*-value=0.9935 for stepwise) were not statistically significant after adjusting for social environmental factors (Figure 2). In these multivariate regression models, the average marginal effects (AMEs) of hot spring were statistically significant for both DED (AME=-0.23, 95% CI=-0.40 -0.07, *p*-value=0.0060) and VHD (AME=0.41, 95% CI=0.17-0.64, *p*-value=0.0008). Moreover, the incorporation of hot spring exposure information enhanced the stepwise logistic regression model performance in DED and VHD prediction (Table S2). These statistic results indicated a robust association between disease (DED and VHD) risk modification and hot spring distribution.

**Figure 2:**
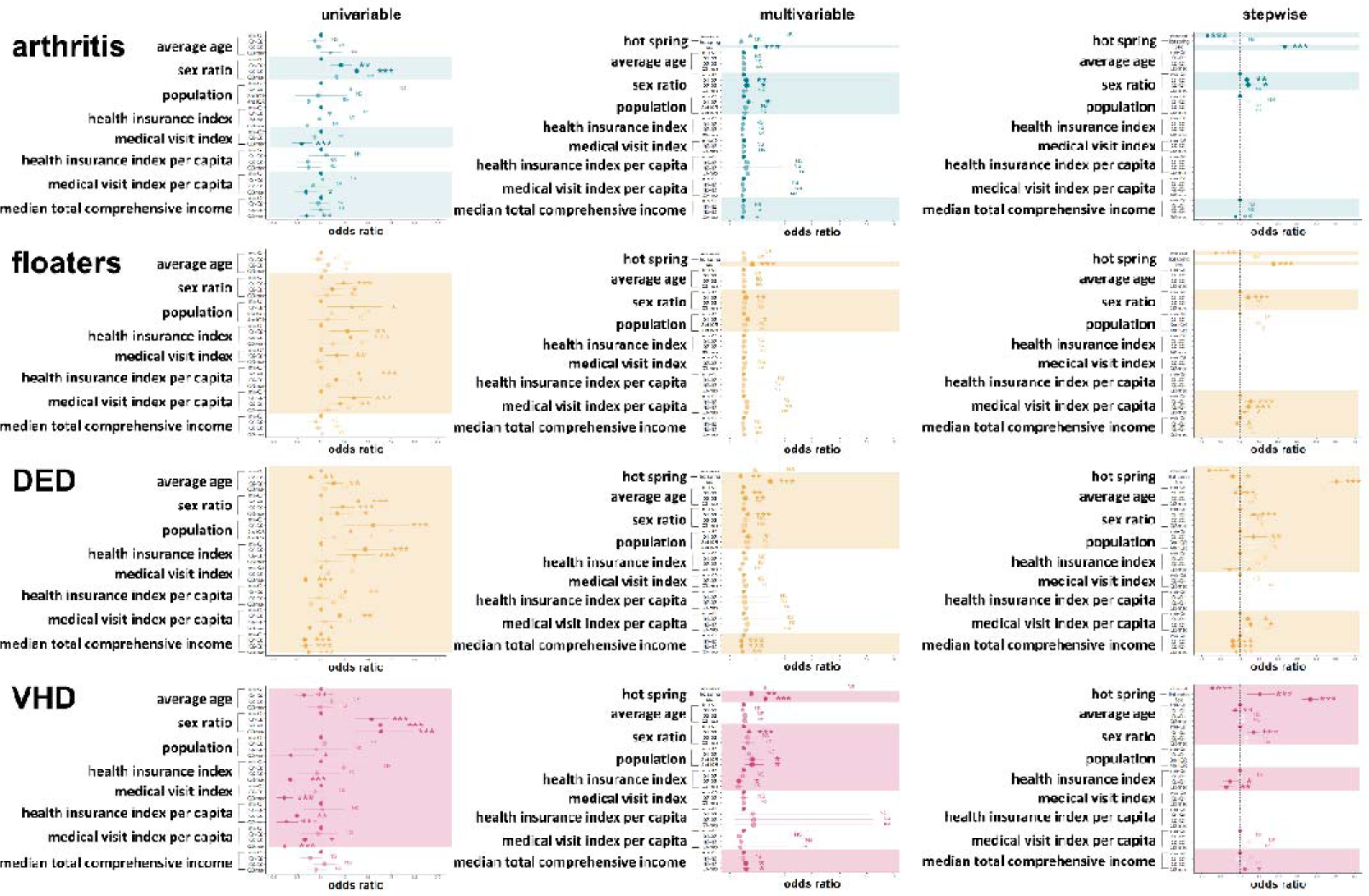
Logistic Regression Models Evaluating Socioeconomic (S) Confounding Effects. The dashed line indicated odds ratio (OR)=1.0 DED, dry eye disease; and VHD, valvular heart disease.

### 3.2. Variant-disease Association Discovery

We applied GWAS (SNP list: Supplementary Table 3-8) and Manhattan plot to visualize the genetic predisposition of the DED and VHD. Interestingly, from the designated subgroup analysis, we noted that the significant (*p*-value<10^-5^) SNPs from non-hot spring subgroup were partial overlay with the parental all participants (for instance, DED: AAK1 rs6755439, rs60177156, rs7131682, rs78027155, rs10438549, *EPIC1* rs62227770, and so on; VHD: rs78714474 and *FOXO3* rs3813498) but were distinct to the hot spring subgroup findings. This SNP discovery discrepancy indicated a potential GxE effect (Figure 3).

**Figure 3:**
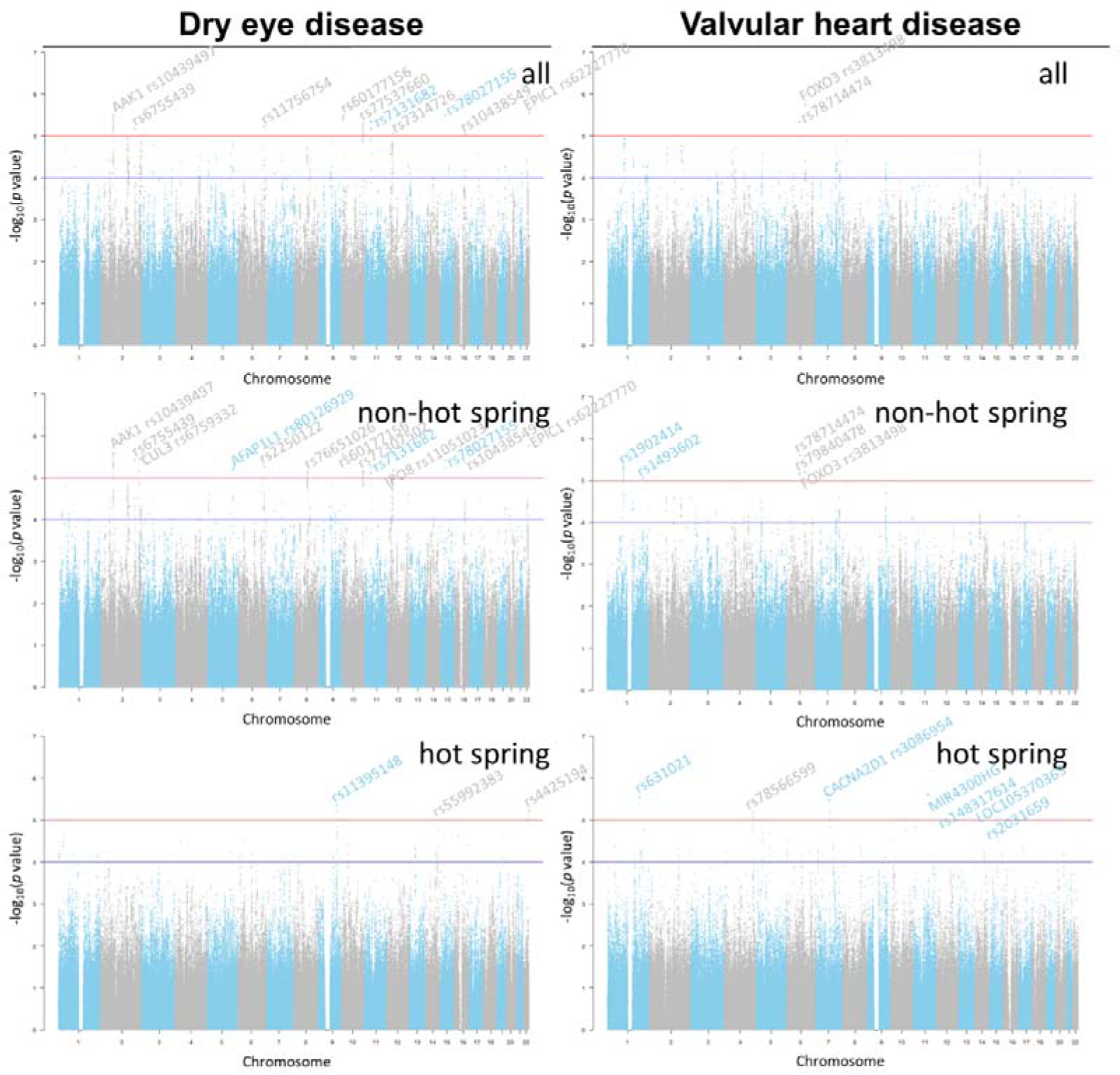
Manhattan Plots of Genome-wide Association Studies (GWASs) among “All”, “Non-hot Spring”, and “Hot Spring” Gene Sets. Blue lines represented *p*-value = 10^-4^ while red lines represented *p*-value=10^-5^.

We then calculated the predict AUC with the derived SNPs list at different significance (Figure 4a). At *p*-value < 0.005, the predict AUC for DED hot spring and VHD hot spring subgroup were 0.92 and 0.82 respectively. However, in the non-hot spring subgroup, the model does not gain AUC increment over the enrolled SNPs.

**Figure 4:**
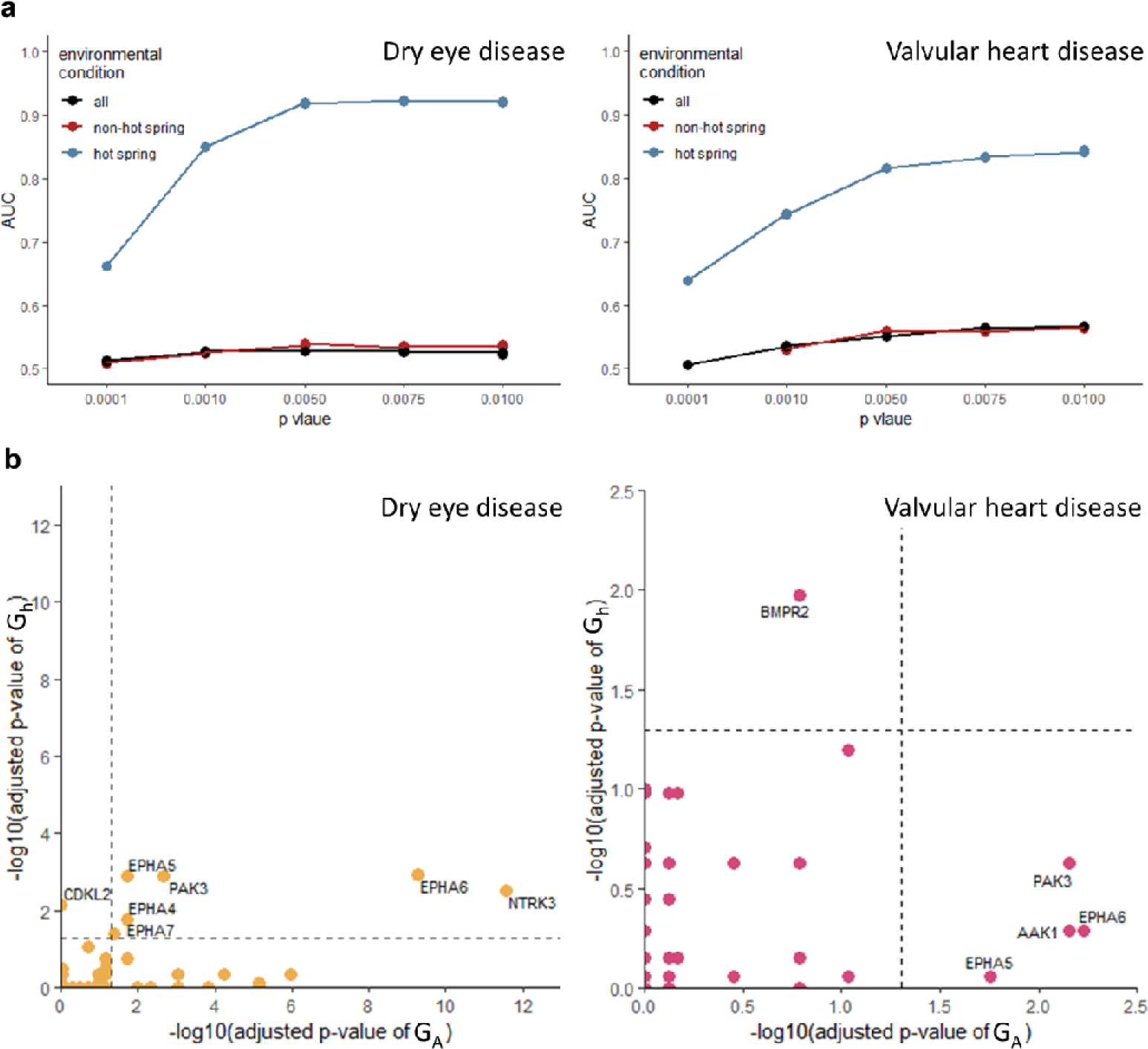
Signaling Pathways of Co-expressed Genes based ARCHS4 Kinases Coexp Library. (a) Screening single nucleotide polymorphisms (SNPs) based on genome-wide significance level threshold (*p*-value< 0.005). AUC, Area Under Curve. (b) Comparing signaling pathways of co-expressed genes between gene set from all population (G_A_) and hot spring-specific gene set (G_h_). The dashed line indicated *p*-value<0.05.

After adjusting LD threshold (r^2^ <0.01) and genome-wide significance threshold (*p*-value <LJ0.005) (Supplementary Figure 2), the VDA results were aggregated and annotated as G_A_, G_N_, and G_H_ for “all”, “non-hot spring”, and “hot spring” subgroup respectively. 73 functional DED SNPs and 19 functional VHD SNPs among G_A_, G_N_, and G_H_ were overlapped (Supplementary Figure 3). After pathway discovery, there were 15 and 4 significant ARCHS4 human kinase pathways in the G_A_ of DED and VHD respectively (adjusted *p*-value<0.05) (Figure 4b). However, CDKL2 kinase pathway was the only significant hit for G_h_ subgroup (Overlapped ratio: 21/299 with *GABARAPL2, C11ORF87, CXADR, DOCK3, ANKRD46, DDX25, PTPRO, ANK2, NALCN, LRP1B, MYO16, TRIM9, PCLO, CACNB4, DLG2, TRIM2, PSD3, TP53BP1, CTNNA2, SRGAP3, CDH18*; OR=2.79, adjusted *p*-value=0.00698). Meanwhile, BMPR2 kinase pathway was the only significantly hit in the VHD G_h_ (Overlapped ratio: 17/299 with *RBFOX1, EPAS1, CACNA2D1, CRIM1, PTPRM, PARVA, ZDHHC21, LAMB1, ATP2B2, KALRN, HIPK2, SIPA1L1, AKT3, EDIL3, CLASP1, KCNH1, OPCML*; OR=3.51, adjusted *p*-value=0.01066).

To investigate the putative molecular interaction in the derived G_h_ hits, we conducted Reactome pathway detection in the PPI after filtering and clustering G_h_ based on protein crosstalk in DED and VHD respectively. Consisting of 158 genes with *RYR2, CACNA1C, TGF*β*1,* and *MAPK8* as four central hub genes, activated pathways in PPI network of DED were related to proposed DED-related pathomechanisms, such as O-linked glycosylation of mucins (R-HSA-913709), Interleukin (IL)-4 and IL-13 signaling (R-HSA-6785807), and interaction between L1 and ankyrins (R-HSA-445095)(Figure 5a and 5b). Consisting of 39 genes with *PARP1, GRM1, NRXN2,* and *GRIA4* as four central hub genes, activated pathways in one of PPI network of VHD were related to proposed VHD-related pathomechanisms, such as Calcium (Ca^2+^) channel opening (R-HSA-112308), YAP1- And WWTR1 (TAZ)-stimulated gene expression (R-HSA-2032785), thrombin signaling (R-HSA-456926), cardiac conduction (R-HSA-5576891),Runt-related transcription factor 2 (Runx-2) (R-HSA-8939243), mitogen-activated protein kinase (MAPK) pathway (R-HSA-5674135), interaction between RAF1 and BRAF (R-HSA-5683057), and signaling by GPCR (R-HSA-112308)(Figure 5c-1 and 5d-1). Comprised 6 genes (*CNOT7, CPEB3, CNOT2, UBXN7, EPAS1,* and *COMMD10*), activated pathways in another of PPI network of VHD were related to proposed VHD-related pathomechanisms, such as autophagy (R-HSA-9612973) and hypoxia (R-HSA-1234174) (Figure 5c-2 and 5d-2).

**Figure 5:**
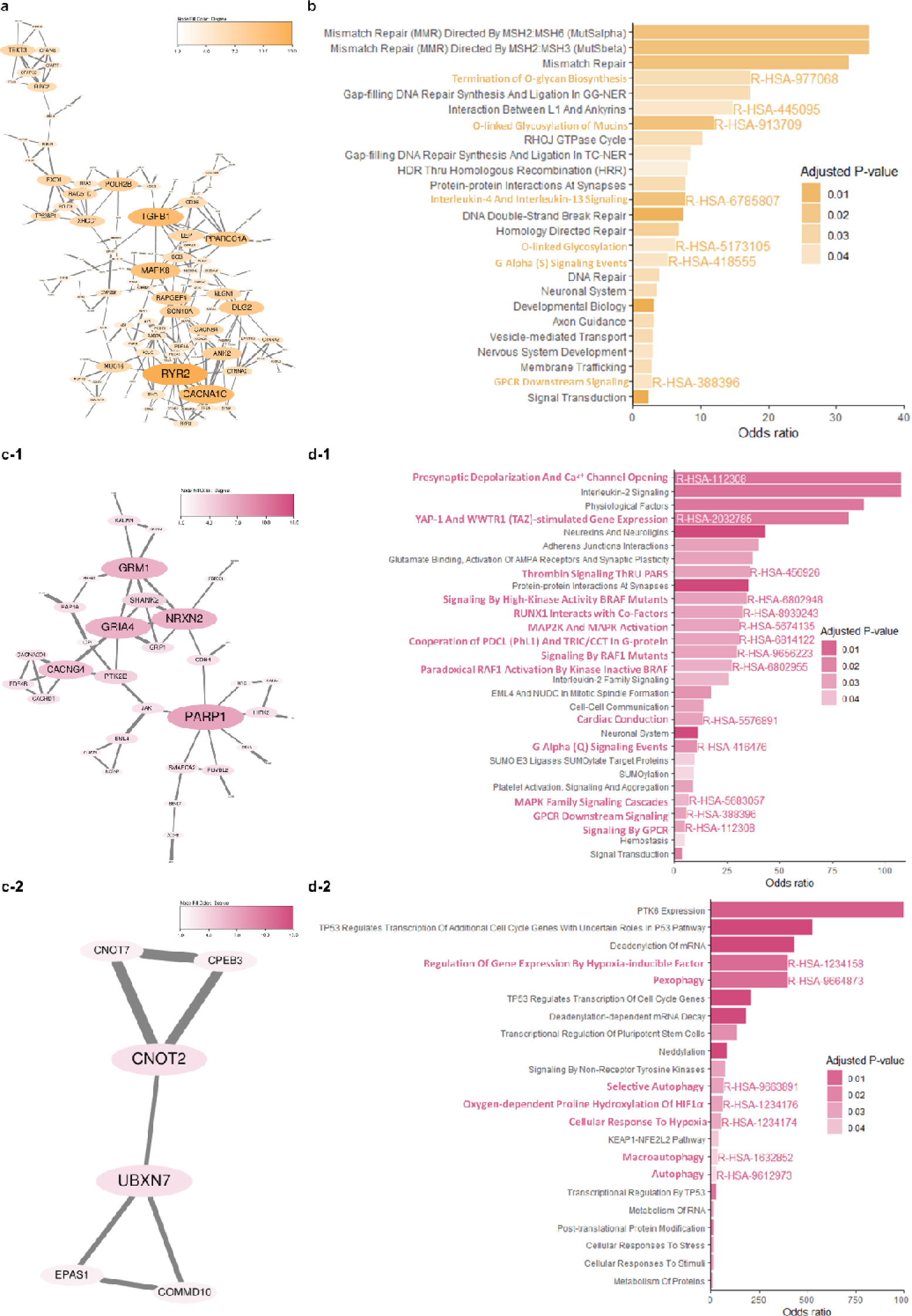
Activated Pathways of Functional Genes within Protein-protein Interaction (PPI) Network of Hot Spring-specific Gene Set (G_h_) based on Reactome 2022. (a) PPI of dry eye disease (DED). (b) Activated pathways of DED. (c) PPI of valvular heart disease (VHD). (d) Activated pathways of VHD. PARS, proteinase activated receptors; HIF1α, hypoxia-inducible factor-1α; MAPK, mitogen-activated protein kinase; and R-HAS, Reactome Homo sapiens.

PRS models were constructed for different environmental conditions (all, non-hot spring, hot spring) based on the associated SNPs discovered from the TWB (G_A_, G_N_, and G_H_). The mean PRS was higher among the cases compared to the controls across all models in both diseases but more significant in G_H_ (AUC: 0.9168 for DED and 0.8153 for VHD) (Figure 6a). With MAF>0.05 and *p*-value of Hardy–Weinberg equilibrium >0.05, SNPs used in PRS model were not rare variants and were independent of environmental conditions (E). However, PRS models with G_H_ could not predict disease risk well in populations without hot spring exposed (AUC: 0.5015 in DED and 0.5066 in VHD), indicating significant GxE relationship. The GxE relationship demonstrated a dose-response effect (Figure 6b). Compared to the participants with PRS in min-Q1, participants with PRS in Q2-Q3 demonstrated 29.11 times odds of developing DED (95% CI=10.36-81.76, *p*-value<0.001) while the individuals with PRS in Q3-max demonstrated 390.81 times odds (95% CI=130.93-1166.52, *p*-value<0.001) (Figure 6c). Compared to the participants with PRS in min-Q1, participants with PRS in Q2-Q3 demonstrated 14.339 times odds of developing VHD (95% CI=6.475-31.754, *p*-value<0.001) while the individuals with PRS in Q3-max demonstrated 34.162 times odds (95% CI=14.842-78.63, *p*-value<0.001) (Figure 6c).

**Figure 6:**
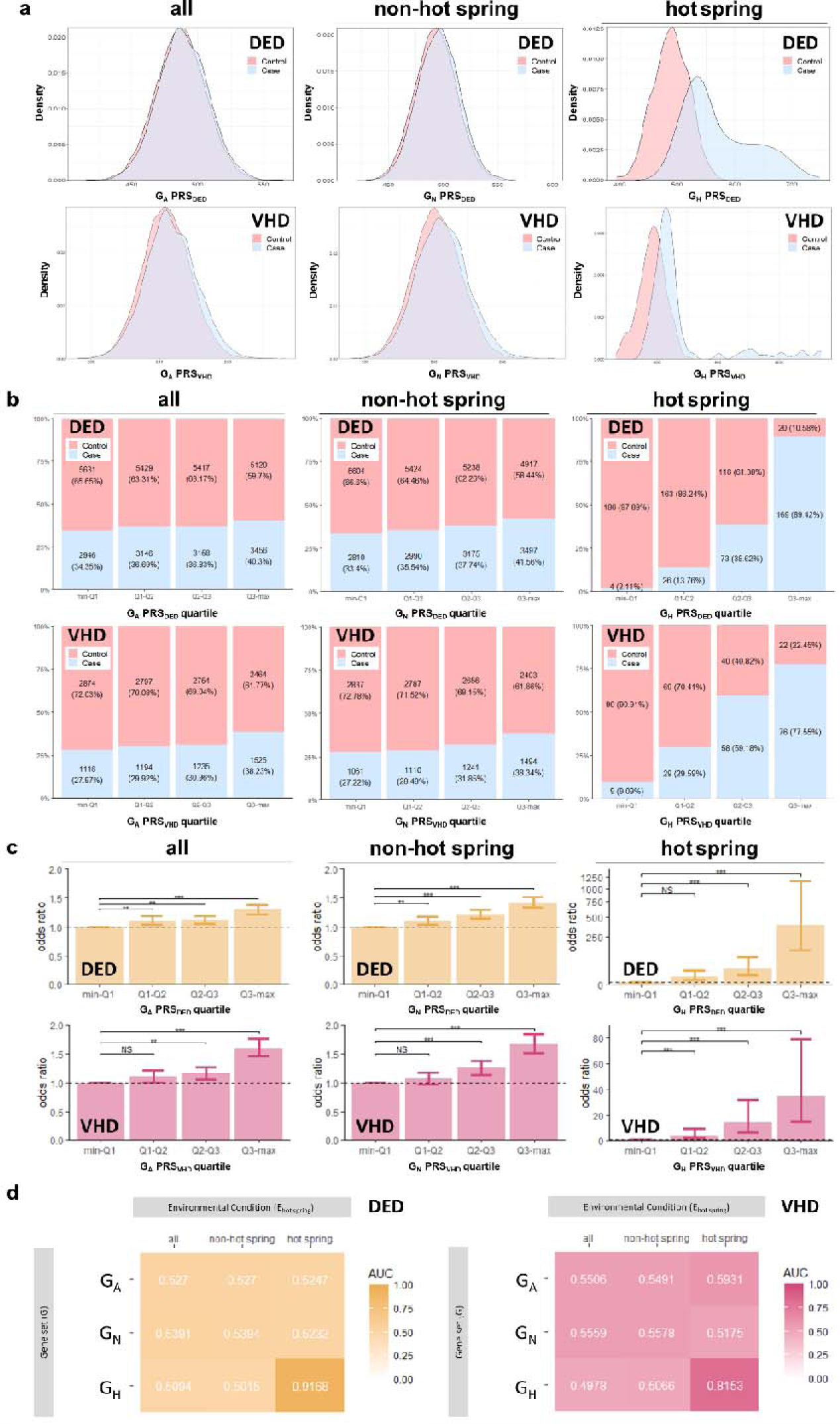
Polygenic Risk Score (PRS) Models Evaluating Gene-environment interactions (GxE). (a) Density distribution among PRS score. After adjusted for linkage disequilibrium (LD) clumping threshold (r^2^<0.01) and genome-wide significance level threshold (*p*-value< 0.005), variant-disease association (VDA) results were depicted as G_A_, G_N_, and G_H_ for “all”, “non hot spring”, and “hot spring” gene sets. (b) Percentage of case and control in each PRS quartile. (c) Odds ratio (OR) of each PRS quartile. The dashed line indicated OR=1.0 (d) Area Under Curve (AUC) of different combinations of gene set (G) and environmental condition (E). DED, dry eye disease; and VHD, valvular heart disease.

### 3.3. GxE Interaction Analysis

While participants with different environmental conditions showed similar density profiles of G_H_PRS (Figure 7a), the disease ratio was significantly different with significant GxE interaction indicated by logistic regression model (*p*-value<10^-16^), showing crossover GxE effect (Figure 7b). Since disease ratio almost remained unchanged across G_H_PRS, GxE effect was calculated based on risk in non-hot spring region as reference. RR tables between different environmental conditions and G_H_PRS indicated hot spring as effect modifier (Figure 7c). Therefore, we enrolled gender and GxE effect to construct DED and VHD predict nomograms (Figure 7d). The DCA curve was performed to demonstrate high clinical net benefit between threshold probability of 20%-48% for DED and 18%-39% for VHD (Figure 7e).

**Figure 7:**
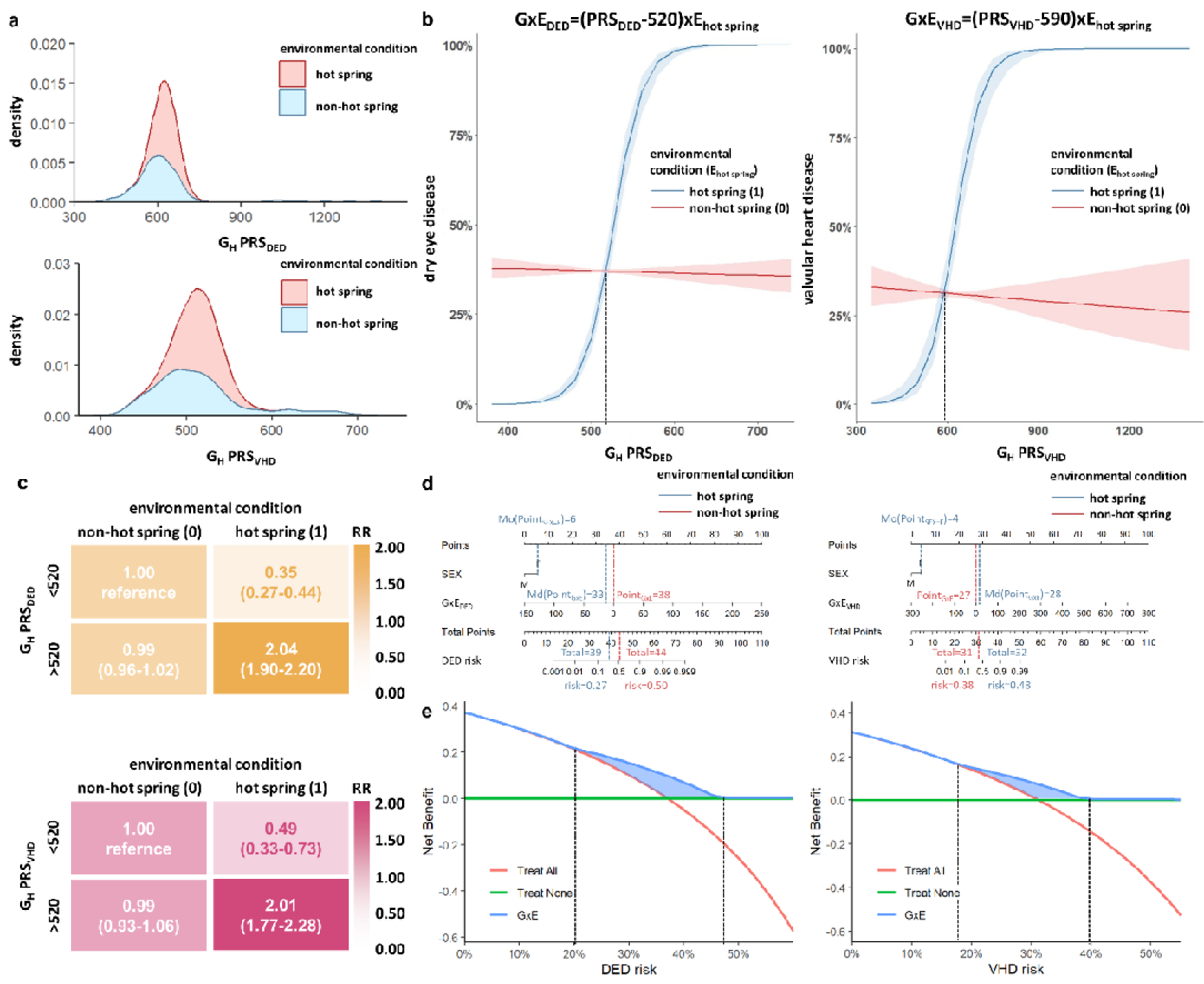
Net Benefit of Identifying the Risk of Gene-environment Interactions (GxE). (a) Density profile of G_H_ polygenic risk score (PRS). (b) Multivariate logistic regression evaluating the GxE effect. (c) Relative risk (RR) with 95% confidence interval (CI) between different environmental conditions and G_H_PRS. (d) Nomogram results estimating predictability of GxE. (e) Decision curve analysis (DCA) assessing net benefit of identifying GxE risk. PRS, polygenic risk score; Mo, mode; Md, median; G_H_, gene set from “hot spring”; DED, dry eye disease; and VHD, valvular heart disease.

## 4. Discussion

By employing multimodal analysis based on genetic data in Taiwan biobank and DA in Opendata platform, we conducted environmental association discovery, variant-disease association discovery, and the GxE interaction analysis to reconstitute GxE effect on DED and VHD. This context dependent GxE interactions were discussed as below:

### 4.1. Hot Spring and Disease Correlation

In the environmental association discovery, residency in hot spring areas was associated with a lower prevalence of dry eye (OR=0.81) and a higher prevalence of valvular heart disease (OR=1.52).

### 4.1.1. Hot Spring-related Factors in DED

Dry eye is a result of increased osmolarity of the tear film and inflammation of the ocular surface (Pflugfelder and de Paiva 2017). It can be influenced by external factors such as desiccating environment and wind exposure (Tsubota et al. 2017). In previous air pollutants studies, higher humidity levels were associated with improved corneal fluorescein staining and tear breakup time (TBUT) (Berg et al. 2020), while participants residing in areas with increased relative humidity were less likely to develop DED (Hwang et al. 2016). Given that hot spring water vapor may increase humidity levels in the immediate vicinity (Nordstrom et al. 2005), it is possible that residing near hot springs were less likely to develop dry eyes. To the best of our knowledge, there are no studies publicly available to study the atmospheric CO_2_ H_2_S and N_2_O influence on DED, future research avenue in the atmospheric (SO_2_, CO_2_ H_2_S, N_2_O) effect on DED occurrence.

### 4.1.2. Hot Spring-related Factors in VHD

Regarding VHD, research has demonstrated that environmental factors, specifically Legionellosis outbreaks in regions such as Taiwan (Hsu et al. 2006) and Japan (ITO et al. 2002), have been linked to the transmission of *Legionella* through hot springs. The Legionella bacteremia often result from pulmonary sources and may cause endocarditis and valvular infections (Brusch 2017). Additionally, other microorganisms, such as *Acanthamoeba spp*. and *Microspora*, have also been associated with hot spring-related transmission and could potentially lead to cardiac complications (Filho et al. 2009). The positive correlation of hot spring distribution and valvular heart disease may be due to ambient pollutants emitted from water or infectious disease spreading through the hydrosphere.

### 4.2. Genetic Effects in DED and VHD

In variant-disease association discovery, we demonstrated that co-expression of kinase pathways (CDKL2 for DED and BMPR2 for VHD) and disease-related reactome pathways were significantly enriched in hot-spring specific functional SNPs.

#### 4.2.1. Hot Spring-specific SNP Genes Co-expressed Kinase Pathway–CDKL2 in DED

Cyclin-dependent kinase-like 2 (CDKL2) belongs to the family of CDC2-related serine/threonine protein kinases. CDKL2 was identified as a target related to Sjögren’s syndrome (Xia et al. 2023) and rheumatoid arthritis (RA) was the most enriched term in the KEGG analysis of CDKL2 methylation geneset (Chen et al. 2021). These studies supported the association between CDKL2 and DED, as dry eye is the most frequent ocular manifestation in patients with Sjögren’s syndrome (>90%) and RA (46.7%) (Kosrirukvongs et al. 2012). The co-expression of CDKL2/CDKL3 (Tracey et al. 2003) and interleukins (IL-15RA, IL-7, IL-1RAPL1, and IL-11) also suggested correlation between CDKL2 and DED as Sjögren’s syndrome, RA, and DED are known to be mediated by cytokine-mediated systemic inflammation. Additionally, CDKL2 expression was detected in DED-involved ocular segments such as the lacrimal gland, cornea, and eyelid, according to the Human Eye Transcriptome Atlas (Tracey et al. 2003). Although a direct link between CDKL2 mutation and DED was not found in current research, we postulate that the effect of CDKL2 on DED is context-dependent and influenced by additional environmental cues.

#### 4.2.2. Activated Pathway in G_h_ SNPs of DED

Activated pathways in the PPI network of DED included O-linked glycosylation of mucins, IL-4 and IL-13 signaling, and interaction between L1 and ankyrins. Transmembrane mucins (such as MUC1, MUC4, and MUC16) (Uchino 2018), large glycoproteins with heavily glycosylated glycans, were revealed to be essential for maintaining ocular surface epithelium lubrication, wettability, and epithelial glycocalyx barrier. In response to inflammatory stimulation of DED, goblet cells and corneal/conjunctival epithelial cells express receptors for inflammatory cytokines such as IL-13 (Baudouin et al. 2019), inducing an important transcriptional factor in epithelial differentiation and goblet cell formation. Activation of transient receptor potential ankyrin 1 (TRPA1) resulted in calcium influx, tear reduction, increased eye blink, and forelimb eye wipe behavior.(Ashok et al. 2023) These G_h_-related pathways implied the underlying pathomechanisms of GxE in DED.

#### 4.2.3. Hot Spring-specific SNP Genes Co-expressed Kinase Pathway–BMPR2 in VHD

Bone morphogenic receptor 2 (*BMPR2*) gene encode a transmembrane serine-threonine kinase receptor that governs SMAD signaling pathways. While SMAD signal cascade was associated with aortic dilation (Balint et al. 2022), dysregulation of the upstream BMPR2 kinase pathways may hold valuable study directions in GxE VHDs. BMPR2 loss of function SNPs were discovered in heritable pulmonary arterial hypertension (PAH) and idiopathic PAH (Chen et al. 2019), these two PAH conditions concurs in 15-60% VHD patients (Magne et al. 2015). Li et al. was the first to report BMPR2 may cause VHD by DNA hypermethylation through down-regulation of *BMPR2* expression (Li et al. 2021). In echo to this report, our result identified BMPR2 pathway as a prominent functional SNP pathway in the hot spring-VHD patients, whereas BMPR2 pathway was not evident in general VHD patients.

#### 4.2.4. Activated Pathway in G_h_ SNPs of VHD

Activated pathways in the PPI network of VHD included Ca^2+^ channel opening, YAP1- And WWTR1 (TAZ)-stimulated gene expression, Runx-2, and MAPK pathway. Recently, combining 2 GWAS studies in 474 and 486 cases from Canada and France, a meta-analysis confirmed the role of RUNX2 and CACNA1C as susceptibility genes of aortic valve stenosis (AS), belonging to the Ca^2+^ signaling pathway (Guauque-Olarte et al. 2015). Moreover, YAP1 has been reported to regulate endothelial to mesenchymal transition (EMT) through modulation of TGFβ-Smad signaling and proliferative activity during cardiac cushion development. MAPK pathway influences calcification, being linked to the expression of a contractile phenotype in valvular interstitial cells. These G_h_-related pathways implied the potential mechanisms of GxE in VHD.

### 4.3. GxE Interaction and Clinical Prospect

In this study, we reported that hot spring residency and underlying polygenetic conjointly modified the disease risk of DED and VHD, whereupon we gathered multi-angle evidence to strengthen the significance of GxE interaction. Although it is generally accepted that both genetic and environmental factors contribute to the development of complex diseases, few GxE examples were replicated, biologically plausible, and methodologically sound interaction with proven clinical relevance and application in daily clinical routines. Therefore, the term “GxE interaction” was often used to express that several factors contribute to disease risk without excluding the possibility of complete independence. To be specific, biological GxE interaction is the joint effect of both genetic and environmental factors that act together in a direct physical or chemical reaction or in the same causal mechanism of disease development (Yang and Khoury 1997) while statistical GxE interaction is defined as “departure from additivity of effects on a specific outcome scale (Rothman and Greenland 1998).” That is, the effect of one factor depends on the level of the other factor. The interaction could be defined as crossover effect or nonremovable interaction only when the interaction remained in any monotone transformation (Thompson 1991). In this study, the interaction of hot spring residency and the level of GH PRS well complied to the requirements of statistical GxE interaction (Figure 7b). However, neither long-term residency effects nor diseases-specific effects have been thoroughly investigated with hot spring. Therefore, we struggled to interpret the biological GxE interaction via GO, co-expressed signaling pathways, and PPI network analysis. Overall, we proposed a novel GxE interaction in hot spring-associated diseases, whereby we suggest further studies are warranted to elucidate the benefits and harms of long-term hot spring residency in human health.

### 4.4. Study Limitation

Despite the favorable results, there are several study limitations in this study. First, the acquisition of VHD and DED disease labels relied on self-reported questionnaires; the definitive diagnosis records and degree of disease severity were intrinsically absent from the biobank data. The hot spring exposure was determined by participant residencies, the exact exposure duration and intensity cannot be estimated and studied from existing accessible data. Although multi-angle evidence supported the GxE interaction in hot spring-related DED and VHD, the concise pathomechanism cannot be inferred from statistical and epidemiological studies. Together, this study served as a pilot study that highlights potential research avenues for GxE effects in hot spring-related diseases.

## 5. Conclusion

Our findings validate the pivotal role of GxE interactions in the epidemiology of DED and VHD within Taiwan’s hot spring regions, underscored by the enhanced disease prediction through PRS models. The study emphasizes the complex interdependencies between genetics and environmental exposures in disease manifestation, reinforcing the need for integrative risk assessments in environmental health studies.

## Data Availability

All data produced in the present study are available upon reasonable request to the authors.

## Statements

### Ethics approval and consent to participate

Ethical approvals were obtained from the Institutional Review Board (IRB) of the Taipei Veterans General hospital with reference numbers 2023-01-006AC. All participants provided written informed consent. This study was conducted in compliance with the Helsinki Declaration.

### Data Sharing

All data is published in this manuscript, the cited manuscripts, or the supplementary appendix. Data can be provided upon request to corresponding authors, and in agreement of terms. The datasets generated and/or analyzed during the current study are available in the Nation-wide Taiwan Biobank1 and Taiwan Biobank2 (https://www.twbiobank.org.tw/), Directorate-General of Budget, Accounting and Statistics (https://eng.dgbas.gov.tw) Ministry of Finance (https://www.mof.gov.tw/), Ministry of Health and Welfare (https://data.gov.tw/dataset/39280, https://data.gov.tw/dataset/39281, https://data.gov.tw/dataset/39282, https://data.gov.tw/dataset/39283) repository.

### Declaration of Interests

The authors declare that they have no competing interest.

### Authors’ Contributions

HYW, KJC and YPY were responsible for the conceptualization and were actively involved in the planning of methodology, investigation, and project administration. PHC and HYT contributed to the formal analysis. HYW, KJC, WC, CYW participated by validation, visualization, writing original draft, reviewing, and editing. YTH, YCW, YCC, CHH, ARH, SHC and CCH provided critical advice. YHC, HJD, CHL, and YCC accessed and verified the underlying data reported in the manuscript. All authors had full access to all the data and responsibility for the decision to submit for publication.

## Acknowledgements

This study is based in part on data from the Big Data Center, Taipei Veterans General Hospital (BDC, TPEVGH). The interpretations and conclusions contained herein do not represent the position of Taipei Veterans General Hospital. The authors thank Department of Statistics, Tamkang University for technological support.

This work was supported by the Taiwan National Science and Technology Council [grant numbers NSTC 112-2321-B-A49-007, NSTC 111-2320-B-A49-028-MY3, NSTC 112-2124-M-038-001, and NSTC 112-2314-B-032-001]; and Taipei Veterans General Hospital [grant number V113C-201, V112C-026 and 112VACS-007].

## Notes

### Competing Interest Statement

The authors have declared no competing interest.

### Funding Statement

This study was funded by the Taiwan National Science and Technology Council [grant numbers NSTC 112-2321-B-A49-007, NSTC 111-2320-B-A49-028-MY3, NSTC 112-2124-M-038-001, and NSTC 112-2314-B-032-001]; and Taipei Veterans General Hospital [grant number V113C-201, V112C-026 and 112VACS-007].

### Author Declarations

IRB of Taipei Veterans General Hospital gave ethical approval for this work

