## Supplementary Table 1 for "Hot Spring Residency and Disease Association: a Crossover Gene-Environment Interaction (GxE) Study in Taiwan"

***Table S1*: Interquartile range of demographic characteristics, economic status (median total comprehensive income), and medical resources of patients living in townships with or without hot spring**

|  | **Township w/ hot spring (n=41)** | **Township w/o hot spring (n=327)** | **p-value** |
| --- | --- | --- | --- |
| **Age** | | | |
| Average (year) | 43.56±3.65 | 43.72±3.18 | 0.770 |
| min-Q1 | 11 (26.8) | 81 (24.8) | 0.309 |
| Q1-Q2 | 6 (14.6) | 86 (26.3) |  |
| Q2-Q3 | 14 (34.1) | 78 (23.9) |  |
| Q3-max | 10 (24.4) | 82 (25.1) |  |
| **Sex ratio** | | | |
| Average | 1.09±0.07 | 1.05±0.10 | 0.003 |
| min-Q1 | 5 (12.2) | 87 (26.6) | 0.007 |
| Q1-Q2 | 5 (12.2) | 87 (26.6) |  |
| Q2-Q3 | 15 (36.6) | 77 (23.5) |  |
| Q3-max | 16 (39.0) | 76 (23.2) |  |
| **Population number** | | | |
| Average | 26667.85±56211.21 | 68140.46±84718.69 | <0.001 |
| min-Q1 | 26 (63.4) | 66 (20.2) | <0.001 |
| Q1-Q2 | 9 (22.0) | 83 (25.4) |  |
| Q2-Q3 | 3 (7.3) | 89 (27.2) |  |
| Q3-max | 3 (7.3) | 89 (27.2) |  |
| **Medical institution** | | | |
| Pharmacy | 6.20±16.71 | 20.58±30.0 | <0.001 |
| Clinic | 9.24±21.67 | 30.92±45.33 | <0.001 |
| Local hospital | 0.24±0.66 | 1.01±1.69 | 0.001 |
| Regional hospital | 0.10±0.49 | 0.23±0.58 | 0.032 |
| Medical center | 0.05±0.22 | 0.06±0.23 | 0.810 |
| **Health insurance index^a^** | | | |
| min-Q1 | 23 (56.1) | 64 (19.6) | <0.001 |
| Q1-Q2 | 10 (24.4) | 85 (26.0) |  |
| Q2-Q3 | 5 (12.2) | 89 (27.2) |  |
| Q3-max | 3 (7.3) | 89 (27.2) |  |
| **Medical visit index^b^** | | | |
| min-Q1 | 23 (56.1) | 64 (19.6) | <0.001 |
| Q1-Q2 | 10 (24.4) | 84 (25.7) |  |
| Q2-Q3 | 5 (12.2) | 90 (27.5) |  |
| Q3-max | 3 (7.3) | 89 (27.2) |  |
| **Health insurance index^a^ per capita** | | | |
| min-Q1 | 14 (34.1) | 78 (23.9) | 0.134 |
| Q1-Q2 | 13 (31.7) | 79 (24.2) |  |
| Q2-Q3 | 9 (22.0) | 83 (25.4) |  |
| Q3-max | 5 (12.2) | 87 (26.6) |  |
| **Medical visit index^b^ per capita** | | | |
| min-Q1 | 14 (34.1) | 78 (23.9) | 0.056 |
| Q1-Q2 | 9 (22.0) | 83 (25.4) |  |
| Q2-Q3 | 14 (34.1) | 78 (23.9) |  |
| Q3-max | 4 (9.8) | 88 (26.9) |  |
| **Median total comprehensive income** | | | |
| Average (k NTD) | 389.59±59.65 | 408.57±70.04 | 0.001 |
| min-Q1 | 11 (26.8) | 80 (24.5) | 0.105 |
| Q1-Q2 | 13 (31.7) | 78 (23.9) |  |
| Q2-Q3 | 13 (31.7) | 80 (24.5) |  |
| Q3-max | 4 (9.8) | 89 (27.2) |  |

**^a^Health insurance index**

= annual expenditure ratio of health insurance E(L) / total number in Taiwan T(L) × number in township N(L, T)

**Medical visit index**

= annual total medical visit ratio A(L) / total number in Taiwan T(L) × number in township N(L, T)

L:levels of care (including clinic, local hospital, regional hospital, medical center) ; T:township

| **L** | **E (%)** | **T** | **A (%)** |
| --- | --- | --- | --- |
| **Clinic** | 19.5 | 10489 | 65.0 |
| **Local hospital** | 13.8 | 339 | 11.0 |
| **Regional hospital** | 26.3 | 80 | 14.0 |
| **Medical center** | 29.1 | 21 | 10.0 |
