## Supplementary Figure 1 for "Hot Spring Residency and Disease Association: a Crossover Gene-Environment Interaction (GxE) Study in Taiwan"

***Figure S1*: Geographic distribution of demographic characteristics, economic status (median total comprehensive income), and medical resources of patients in Taiwan**

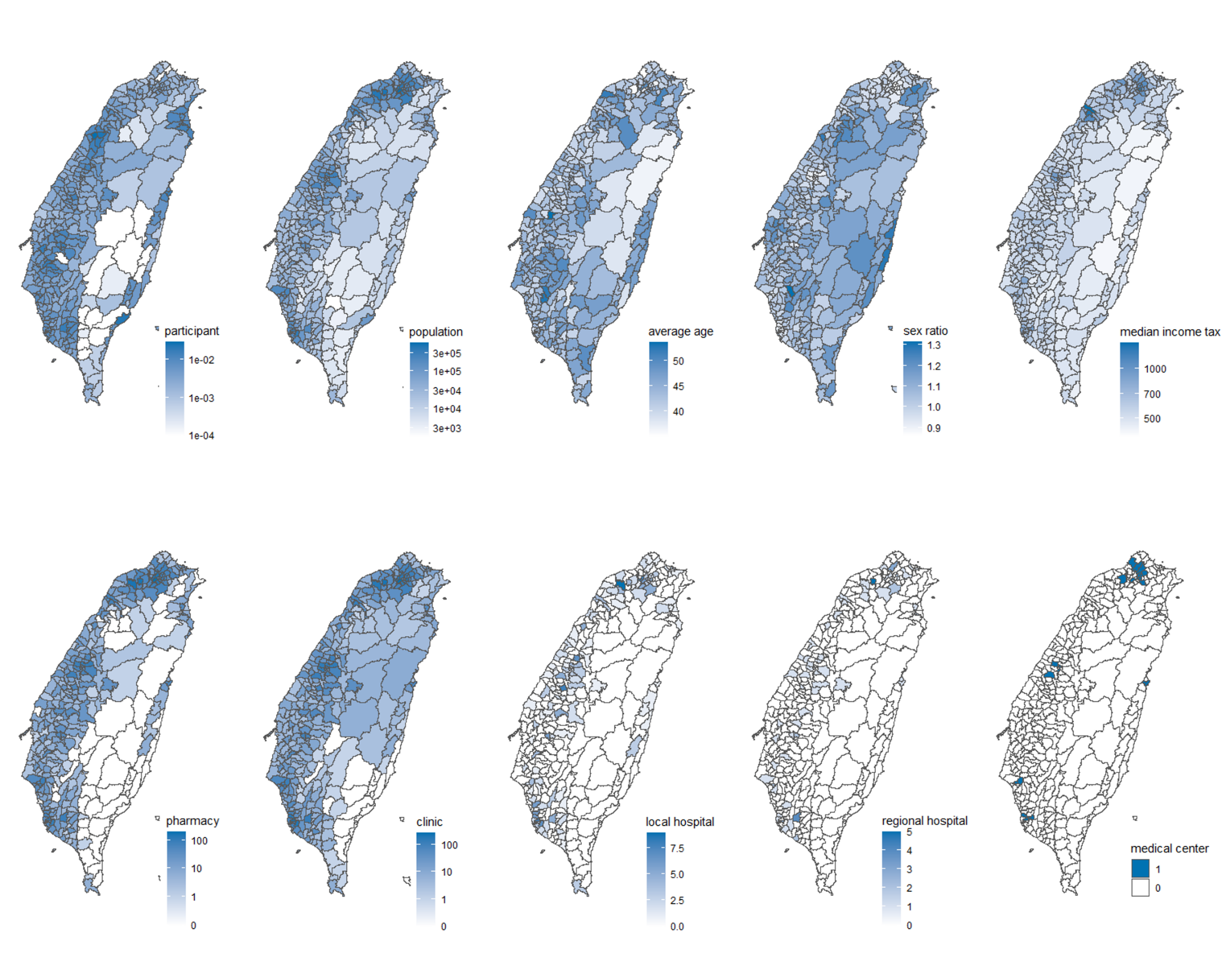
