## Supplementary Table 2 for "Hot Spring Residency and Disease Association: a Crossover Gene-Environment Interaction (GxE) Study in Taiwan"

***Table S2*: Predictive performance of various stepwise regression models compared between hot spring and non-hot spring regions after adjusting social factors**

| **Disease** | **Region** | **AIC** | **AUC** | **Deviance** | **R^2^** |
| --- | --- | --- | --- | --- | --- |
| **Arthritis** | non hot spring | 28251 | 0.5986* | 28228 | 0.022 |
|  | hot spring | 28250* | 0.5986 | 28226* | 0.022 |
| **Floaters** | non hot spring | 46124* | 0.5847 | 46095 | 0.0169 |
|  | hot spring | 46126 | 0.5847 | 46095 | 0.0169 |
| **DED** | non hot spring | 42599 | 0.6533 | 42554 | 0.0587 |
|  | hot spring | 42595* | 0.6533* | 42549* | 0.0589* |
| **VHD** | non hot spring | 19065 | 0.6435 | 19030 | 0.0461 |
|  | hot spring | 19055* | 0.6441* | 19018* | 0.0467* |

*Better predictive performance, AIC: Akaike information criterion, AUC: Area Under Curve
