## Supplementary Figure 2 for "Hot Spring Residency and Disease Association: a Crossover Gene-Environment Interaction (GxE) Study in Taiwan"

***Figure S2:* Functional SNPs defined by the threshold determination of pairwise squared correlation (r^2^) and genome-wide significance level (p value) in all, non-hot spring, and hot spring populations**
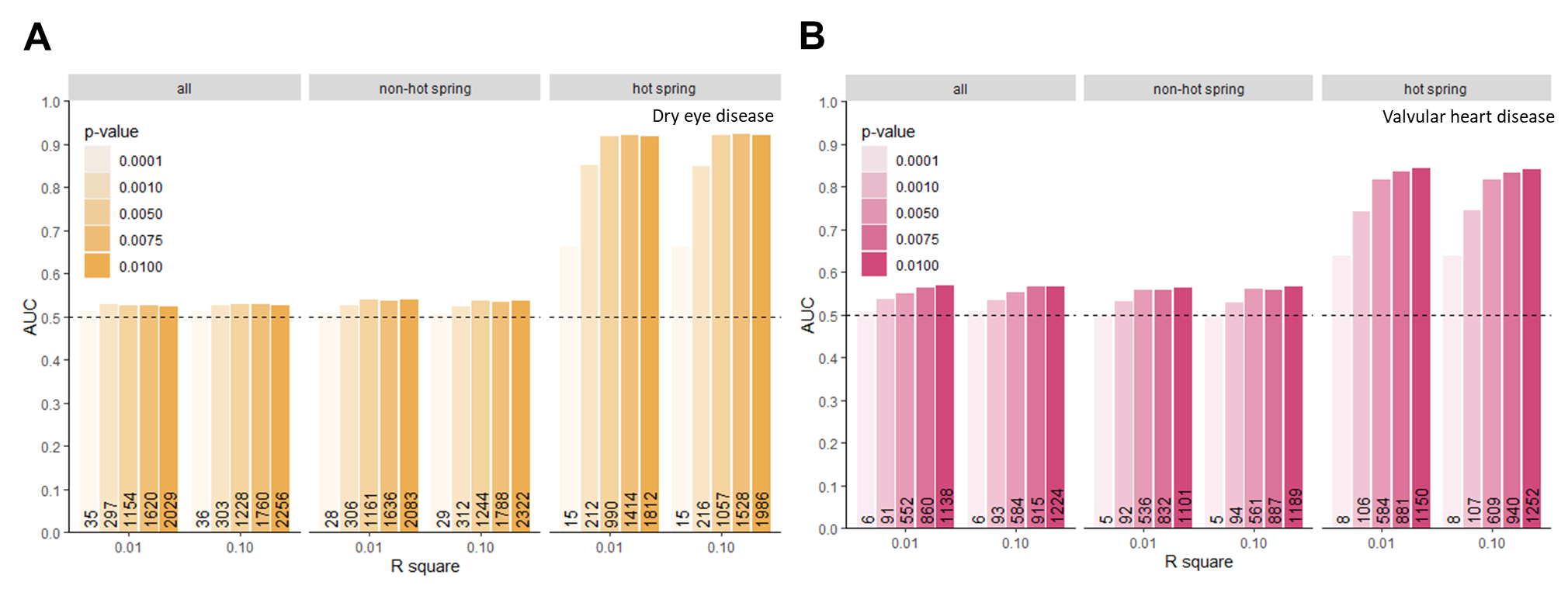
