## Supplementary Figure 3 for "Hot Spring Residency and Disease Association: a Crossover Gene-Environment Interaction (GxE) Study in Taiwan"

***Figure S3:* Venn diagram of functional SNP genes from all, non-hot spring, and hot spring population**

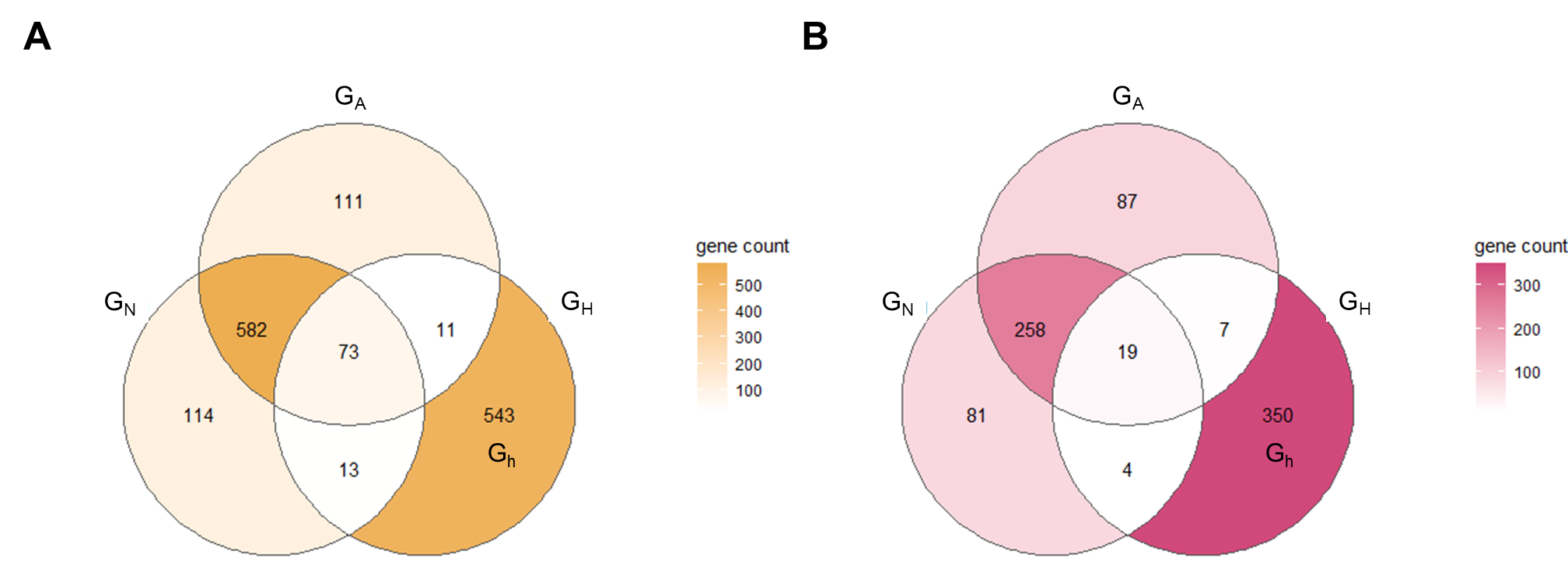

A: DED

B: VHD

G_A_: all population gene sets

G_N_: non-hot spring population gene sets

G_H_: hot spring population gene sets

G_h_: G_H_ that did not overlap with G_A_ and G_N_
