## Supplementary Table 3 for "Hot Spring Residency and Disease Association: a Crossover Gene-Environment Interaction (GxE) Study in Taiwan"

***Table S3:* DED related SNPs of GWAS and their corresponding functional genes in G_A_**

| **Chr** | **SNP** | **bp** | **Allele** | **OR** | **p-value** | | **Gene** |
| --- | --- | --- | --- | --- | --- | --- | --- |
| 15 | rs79614527 | 47881984 | G | 0.8638 | 1.552 x 10 | -6 | - |
| 10 | rs1934358 | 9922745 | C | 1.106 | 5.241 x 10 | -6 | - |
| 11 | rs379388 | 17922699 | T | 0.9048 | 5.669 x 10 | -6 | SERGEF |
| 22 | rs62227770 | 47669099 | T | 0.8829 | 5.899 x 10 | -6 | EPIC1 |
| 11 | rs10743083 | 8614803 | A | 1.113 | 7.020 x 10 | -6 | TRIM66 |
| 1 | rs12031839 | 33921109 | G | 1.213 | 1.981 x 10 | -5 | CSMD2 |
| 10 | rs11592650 | 18343996 | G | 0.8772 | 1.991 x 10 | -5 | CACNB2 |
| 11 | rs4936032 | 112331499 | A | 0.8204 | 2.049 x 10 | -5 | LINC02762 |
| 1 | rs61771304 | 76041188 | A | 1.213 | 2.300 x 10 | -5 | - |
| 9 | rs1402792 | 28599506 | G | 0.9075 | 2.354 x 10 | -5 | LINGO2 |
| 6 | rs3020314 | 151949537 | T | 1.12 | 2.429 x 10 | -5 | ESR1 |
| 11 | rs263101 | 35940699 | C | 1.109 | 2.522 x 10 | -5 | LDLRAD3 |
| 10 | rs10749457 | 122284546 | A | 0.8769 | 3.461 x 10 | -5 | BTBD16 |
| 2 | rs6755439 | 156380227 | A | 0.9032 | 3.505 x 10 | -5 | - |
| 10 | rs72779330 | 43026867 | G | 0.8336 | 3.749 x 10 | -5 | - |
| 1 | rs35655955 | 213690222 | G | 0.8661 | 5.222 x 10 | -5 | - |
| 1 | rs613158 | 54220791 | C | 1.089 | 5.226 x 10 | -5 | SSBP3 |
| 5 | rs13354337 | 72299775 | A | 1.089 | 5.328 x 10 | -5 | MRPS27 |
| 12 | rs1148690 | 90357385 | C | 1.123 | 5.369 x 10 | -5 | - |
| 1 | rs643535 | 84040825 | A | 1.136 | 6.018 x 10 | -5 | - |
| 8 | rs13266927 | 82135890 | G | 1.089 | 6.240 x 10 | -5 | LINC02839 |
| 9 | rs35906092 | 90088786 | CTCA | 1.101 | 6.652 x 10 | -5 | - |
| 7 | rs73332293 | 35510392 | T | 1.149 | 7.167 x 10 | -5 | - |
| 5 | rs962435 | 34621746 | A | 0.9205 | 7.329 x 10 | -5 | - |
| 2 | rs78131129 | 67247142 | A | 1.136 | 7.541 x 10 | -5 | LINC01828 |
| 6 | rs12523801 | 40463835 | T | 0.8549 | 8.141 x 10 | -5 | LRFN2 |
| 20 | rs74674215 | 18378258 | A | 0.8625 | 8.450 x 10 | -5 | LINC00851 |
| 9 | rs4741817 | 3197892 | C | 0.9211 | 8.559 x 10 | -5 | LINC01231 |
| 1 | rs9425301 | 172394675 | A | 0.8859 | 8.821 x 10 | -5 | DNM3 |
| 1 | rs565436 | 55058928 | G | 0.8753 | 8.984 x 10 | -5 | PCSK9 |
| 13 | rs72662043 | 109437167 | T | 1.14 | 8.987 x 10 | -5 | - |
| 16 | rs10438549 | 60581163 | G | 1.084 | 9.416 x 10 | -5 | - |
| 9 | rs79617032 | 95775314 | A | 1.153 | 9.574 x 10 | -5 | ERCC6L2AS1 |
| 20 | rs6064411 | 56542963 | C | 1.085 | 9.659 x 10 | -5 | - |
| 17 | rs9895250 | 9405949 | A | 1.14 | 9.979 x 10 | -5 | STX8 |
