## Supplementary Table 4 for "Hot Spring Residency and Disease Association: a Crossover Gene-Environment Interaction (GxE) Study in Taiwan"

***Table S4:* DED related SNPs of GWAS and their corresponding functional genes in G_N_**

| **Chr** | **SNP** | **bp** | **Allele** | **OR** | **p-value** | | **Gene** |
| --- | --- | --- | --- | --- | --- | --- | --- |
| 15 | rs79614527 | 47881984 | G | 0.8645 | 2.219 x 10 | -6 | - |
| 22 | rs62227770 | 47669099 | T | 0.8796 | 3.825 x 10 | -6 | EPIC1 |
| 10 | rs1934358 | 9922745 | C | 1.104 | 1.001 x 10 | -5 | - |
| 1 | rs12031839 | 33921109 | G | 1.223 | 1.007 x 10 | -5 | CSMD2 |
| 11 | rs263101 | 35940699 | C | 1.114 | 1.332 x 10 | -5 | LDLRAD3 |
| 6 | rs3020314 | 151949537 | T | 1.124 | 1.599 x 10 | -5 | ESR1 |
| 11 | rs379388 | 17922699 | T | 0.9085 | 1.612 x 10 | -5 | SERGEF |
| 1 | rs61771304 | 76041188 | A | 1.215 | 2.197 x 10 | -5 | - |
| 11 | rs10743083 | 8614803 | A | 1.107 | 2.333 x 10 | -5 | TRIM66 |
| 10 | rs11592650 | 18343996 | G | 0.8794 | 3.330 x 10 | -5 | CACNB2 |
| 11 | rs4936032 | 112331499 | A | 0.8245 | 3.807 x 10 | -5 | LINC02762 |
| 9 | rs1402792 | 28599506 | G | 0.9098 | 4.566 x 10 | -5 | LINGO2, KCTD10P1 |
| 10 | rs72779330 | 43026867 | G | 0.8343 | 4.641 x 10 | -5 | - |
| 1 | rs643535 | 84040825 | A | 1.139 | 4.895 x 10 | -5 | - |
| 1 | rs613158 | 54220791 | C | 1.09 | 5.220 x 10 | -5 | SSBP3 |
| 8 | rs13266927 | 82135890 | G | 1.09 | 5.904 x 10 | -5 | LINC02839 |
| 16 | rs10438549 | 60581163 | G | 1.088 | 6.037 x 10 | -5 | - |
| 1 | rs565436 | 55058928 | G | 0.8713 | 6.086 x 10 | -5 | PCSK9 |
| 5 | rs3822407 | 180592341 | A | 1.156 | 6.350 x 10 | -5 | SCGB3A1 |
| 4 | rs4694549 | 72818107 | C | 1.092 | 6.894 x 10 | -5 | - |
| 9 | rs79617032 | 95775314 | A | 1.158 | 6.951 x 10 | -5 | ERCC6L2AS1 |
| 2 | rs6755439 | 156380227 | A | 0.9061 | 7.025 x 10 | -5 | - |
| 10 | rs10749457 | 122284546 | A | 0.8807 | 7.167 x 10 | -5 | BTBD16 |
| 2 | rs76645968 | 53600549 | C | 1.184 | 8.400 x 10 | -5 | ASB3 |
| 6 | rs707827 | 15327269 | A | 0.922 | 9.558 x 10 | -5 | JARID2, RNU6645P |
| 5 | rs13354337 | 72299775 | A | 1.087 | 9.608 x 10 | -5 | MRPS27 |
