## Supplementary Table 5 for "Hot Spring Residency and Disease Association: a Crossover Gene-Environment Interaction (GxE) Study in Taiwan"

***Table S5:* DED related SNPs of GWAS and their corresponding functional genes in G_H_**

| **Chr** | **SNP** | **bp** | **Allele** | **OR** | ***p-value*** | | **Gene** |
| --- | --- | --- | --- | --- | --- | --- | --- |
| 13 | rs56664582 | 80444923 | G | 4.024 | 5.628 x 10 | -6 | - |
| 5 | rs116555818 | 75305109 | A | 2.086 | 1.592 x 10 | -5 | - |
| 6 | rs34103781 | 123794523 | CTG | 0.5199 | 2.603 x 10 | -5 | - |
| 3 | rs1512907 | 89306811 | C | 2.002 | 5.113 x 10 | -5 | EPHA3 |
| 10 | rs57197814 | 52290545 | T | 3.651 | 5.270 x 10 | -5 | PRKG1 |
| 4 | rs150269442 | 164080684 | G | 3.799 | 5.572 x 10 | -5 | MARCHF1 |
| 19 | rs76384439 | 43547432 | T | 2 | 5.810 x 10 | -5 | XRCC1 |
| 7 | rs12539905 | 10926702 | T | 2.988 | 5.876 x 10 | -5 | - |
| 8 | rs7842670 | 134904206 | G | 3.156 | 6.976 x 10 | -5 | - |
| 18 | rs9966547 | 61789282 | C | 0.3518 | 7.165 x 10 | -5 | - |
| 2 | rs72958376 | 99512541 | G | 2.857 | 7.208 x 10 | -5 | - |
| 14 | rs75401508 | 79639067 | C | 1.952 | 7.279 x 10 | -5 | NRXN3 |
| 2 | rs13017411 | 122364669 | T | 2.177 | 7.508 x 10 | -5 | - |
| 1 | rs11208080 | 62986405 | G | 0.5479 | 8.060 x 10 | -5 | LINC01739 |
| 1 | rs2618652 | 237554482 | A | 0.5553 | 9.138 x 10 | -5 | RYR2 |

***Table S6:* VHD related SNPs of GWAS and their corresponding functional genes in G_A_**

| **Chr** | **SNP** | **bp** | **Allele** | **OR** | **p-value** |  | **Gene** |
| --- | --- | --- | --- | --- | --- | --- | --- |
| 12 | rs1860371 | 46791488 | C | 0.7814 | 1.006 x 10 | -5 | - |
| 18 | rs1433859 | 40347065 | C | 1.517 | 1.360 x 10 | -5 | USP45 |
| 6 | rs10457650 | 99468306 | A | 1.297 | 5.417 x 10 | -5 | LINC02112 |
| 5 | rs1005160 | 9796947 | C | 0.7829 | 6.360 x 10 | -5 | - |
| 5 | rs72802041 | 123937142 | T | 1.379 | 9.762 x 10 | -5 | HABP2 |
| 10 | rs11575684 | 113578856 | G | 1.249 | 9.855 x 10 | -5 | ARHGAP26 |

***Table S7:* VHD related SNPs of GWAS and their corresponding functional genes in G_N_**

| **Chr** | **SNP** | **bp** | **Allele** | **OR** | **p-value** |  | **Gene** |
| --- | --- | --- | --- | --- | --- | --- | --- |
| 12 | rs1860371 | 46791488 | C | 0.7677 | 3.376 x 10 | -6 | LINC00499 |
| 4 | rs6825529 | 138572300 | T | 0.7917 | 3.250 x 10 | -5 | USP45 |
| 6 | rs10457650 | 99468306 | A | 1.304 | 5.012 x 10 | -5 | ARHGAP26 |
| 5 | rs258796 | 143232923 | A | 1.277 | 7.248 x 10 | -5 | LINC02112 |
| 5 | rs1005160 | 9796947 | C | 0.7814 | 7.579 x 10 | -5 | - |

***Table S8:*VHD related SNPs of GWAS and their corresponding functional genes in G_H_**

| **Chr** | **SNP** | **bp** | **Allele** | **OR** | ***p-value*** | | **Gene** |
| --- | --- | --- | --- | --- | --- | --- | --- |
| 3 | rs13318975 | 153111491 | C | 5.782 | 2.778 x 10 | -5 | - |
| 18 | rs12454358 | 35520961 | G | 8.849 | 3.792 x 10 | -5 | ARRB1 |
| 11 | rs7938332 | 75313198 | T | 4.893 | 3.896 x 10 | -5 | LINC00448 |
| 13 | rs9317256 | 62774648 | T | 7.03 | 5.748 x 10 | -5 | PDE11A |
| 2 | rs79361463 | 177991642 | A | 6.837 | 7.308 x 10 | -5 | ARHGAP22 |
| 10 | rs57601353 | 48521137 | T | 6.712 | 7.868 x 10 | -5 | JAK1 |
| 1 | rs74889977 | 64964244 | G | 4.197 | 7.941 x 10 | -5 | - |
| 9 | rs13292166 | 12111416 | A | 5.127 | 8.091 x 10 | -5 | ZDHHC21 |
